# Impact of prolonged video viewing on physical and mental stress: An analysis of autonomic nervous system activity and salivary stress markers

**DOI:** 10.1101/2025.02.04.25321652

**Authors:** Hiroki Matsui, Rina Tanabe, Rikako Mogi, Yuka Sugiura, Hana Yokomachi, Tomoyuki Yokoyama

## Abstract

In recent years, online lectures and conferences have become widespread, but problems such as stress and adjustment disorders caused by watching long videos have become a problem. Few studies have objectively evaluated the minute changes in stress associated with such prolonged viewing. This study aimed to determine the effects of prolonged video viewing on physical and mental stress.

Twenty-six students (10 males and 16 females, aged 21–24) from University A participated in this study. The subjects watched both a high-interest and a low-interest video for 90 minutes each and then subjectively evaluated the video content. Heart rate variability analysis was used to assess heart rate and autonomic nervous system changes during video viewing and saliva samples were collected before, during, and after viewing the videos to measure the concentration of stress markers in the saliva. This study was approved by the Gunma University Medical Ethics Review Committee for Human Subjects.

The results of the subjective evaluation showed that scores for concentration, comprehension, and interest were significantly higher, while the fatigue score was lower for the high-interest video than for the low-interest video. Both videos caused a decrease in heart rate and low sympathetic nerve activity, and an increase in parasympathetic nerve activity; however, the changes were more pronounced with the high-interest video. In addition, the differences in subjective scores between the high- and low-interest videos were more pronounced in women, with salivary chromogranin A also showing opposite trends for the two videos.

This study demonstrates that watching two different types of videos for extended periods affects autonomic nervous system activity, heart rate, and stress hormones in saliva. Since trends differ depending on the type of video and the participant’s sex, attention should be paid to the video content and the individual subject when examining stress during video viewing.

## Introduction

The spread of SARS-CoV-2 infection has led to a rapid increase in remote working and remote learning systems. These environmental changes have led to health problems, such as adverse effects on sleep, stress, depression, and adjustment disorders.[1, 2] In particular, there have been reports of adverse effects on sleep onset and quality among young people and those who do not use devices.[3] In addition, remote learning using devices has become widespread during the COVID-19 pandemic, with some cases using devices for more than five hours per day. Results from a cohort study indicate that screen time and smartphone use are positively correlated with academic stress scores and are part of academic stress, such as increased mental health problems.[4, 5] However, the impact of watching long videos using such devices on mental health and the effects on physical problems associated with mental stress have not been examined.

The autonomic nervous system and sympathetic and parasympathetic nervous systems are involved in fatigue- and stress-induced changes in the body. Sympathetic and parasympathetic activity can be measured noninvasively by spectral analysis of heart rate variability, which is composed of low-frequency (LF) and high-frequency (HF) components. The high-frequency component (HF) is parasympathetic, and the low-frequency to high-frequency power values (LF/HF) ratio indicates sympathetic activity. Changes in activity are associated with depressive symptoms and adjustment disorders.[6, 7] Teenagers with addictive online behaviors have also been shown to have increased LF/HF and decreased HF when exposed to visual information.[8] High sympathetic activity has also been reported to cause sleep disturbances and is an essential indicator of the degree of stress associated with digital devices.[9]

When humans are exposed to stress, two types of responses occur: an endocrine response in the hypothalamic-pituitary-adrenocortical (HPA) axis and an autonomic response in the sympathetic-adrenomedullary (SAM) system.[10] Cortisol is secreted from the adrenal cortex into circulating plasma via the hypothalamic-pituitary axis as an acute response to stress. Cortisol is transferred from the plasma to the saliva and is measured as a free hormone independent of salivary flow.[11] Salivary cortisol increases in response to mental stress and is used as a stress biomarker.[12, 13] Salivary glands secrete salivary amylase, which is modulated by sympathetic and parasympathetic nerves.[14] Acute mental stress can activate the SAM system and induce a short period of amylase secretion from the salivary glands.[15, 16] The salivary glands also produce chromogranin A (CgA) in response to the stimulation of the autonomic nervous system.[17] The salivary CgA is also an indicator of SAM system hyperactivity and has been reported to increase rapidly upon acute mental stress.[18, 19] Mental stress comprises multiple dimensions. Different systems interact over different time courses, necessitating the simultaneous evaluation of multiple salivary markers to obtain a whole picture.[20]

The effects of stress caused by prolonged screen viewing on the sympathetic and parasympathetic nervous systems and multiple salivary hormones are yet to be clarified. In this study, we examined whether changes in stress caused by prolonged viewing of two different types of videos in students could be detected by measuring autonomic nervous system and salivary stress markers.

## Materials and Methods

### Participants and measurement protocol

The study was conducted with 26 university students (10 males and 16 females) aged 21 to 24 years (22.1 ± 0.7 years). Participants were recruited from January 10th to December 31st, 2021. When recruiting participants, we explained the study outline and obtained voluntary written consent before participation.

The measurement protocol is illustrated in Fig 1. Before watching the video, each subject completed a questionnaire regarding their current and previous medical history, video-viewing habits, and physical condition on the day of the study. The subjects were fitted with electrodes for heart rate variability measurement, started the measurement, and rested for 10 minutes. The participants watched two types of videos for 90 minutes. The two types of videos used were those of high interest (comedy movies) and those of low interest (lecture videos in a field different from the participant’s major). Several students of the same generation who were not participants selected each video as the subject of this study. The same participants watched the high- and low-interest videos at the same time of day on different days at random and were measured in the same manner. After viewing each video for 90 minutes, participants rated their interest, concentration, understanding of the content, and fatigue using a 10-point (1–10) Visual Analog Scale (VAS).

**Fig 1.**
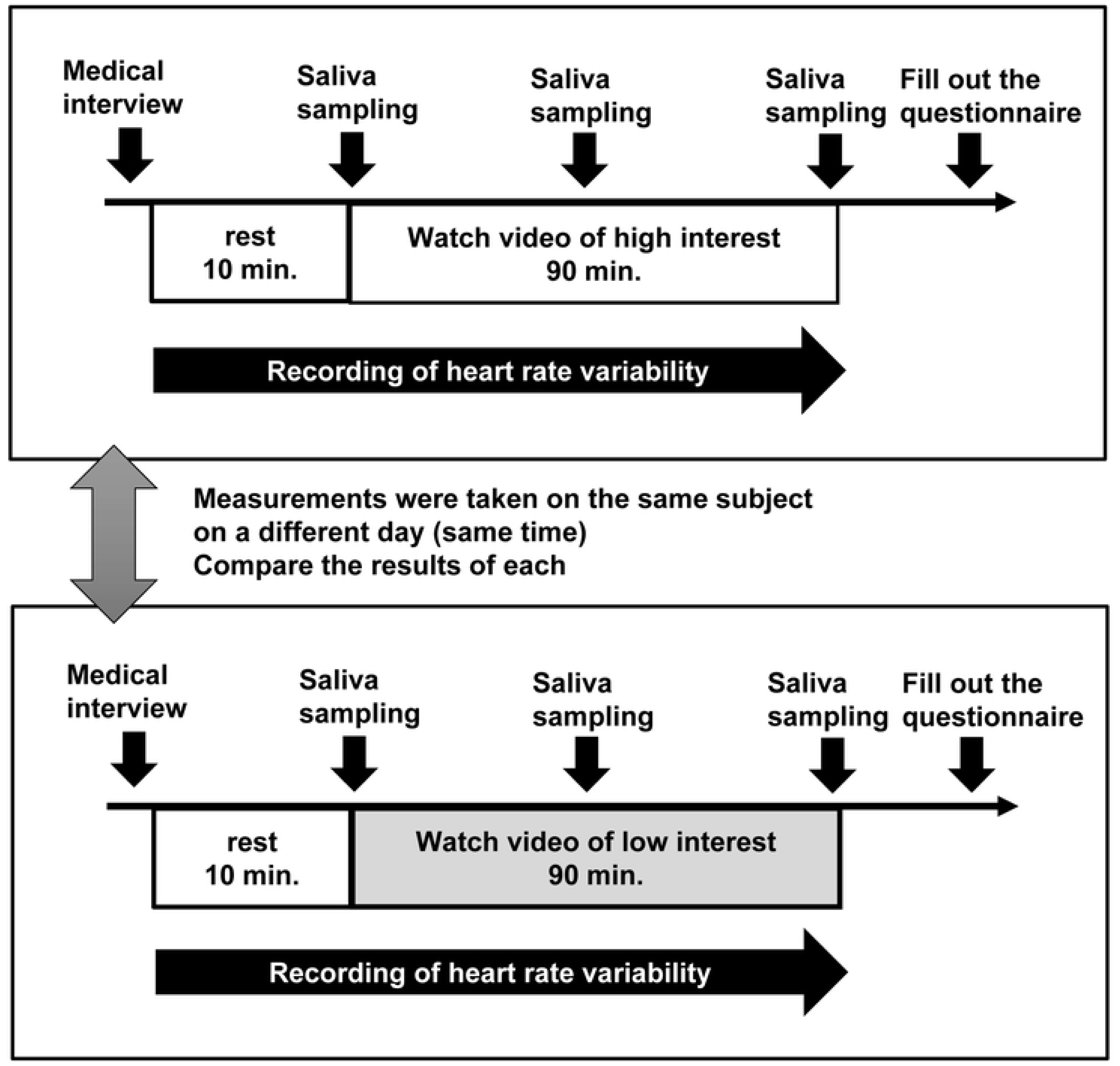
Study design for viewing videos of high interest and low interest. In each session, subjects watched a 90-minute video for 10 minutes after the interview and then answered a questionnaire about the video. Down arrows indicate the interview timing, questionnaire responses, and saliva sampling. Rightward arrows indicate the measurement of heart rate and heart rate variability.

A real-time heart rate variability analyzer measured changes in autonomic nervous activity during video viewing (MemCalc/Bonaly Light, GMS, Japan). Frequency analysis of the heartbeats obtained from the RR interval was analyzed using autoregressive power spectrum analysis applied to the time series. Low-frequency (0.04-0.15 Hz, Low-Frequency: LF) and high-frequency (0.15-0.4 Hz, High-frequency: HF) spectral bands were identified, and the area under each peak was calculated in absolute units (ms^2^). HF was used as an index of cardiac parasympathetic nervous system activity, and LF/HF was used as an index of the cardiac sympathetic nervous system activity.

Saliva samples were collected at three time points: immediately before watching the video (first time), 45 minutes after the start of watching the video (second time), and 90 minutes after watching the video (third time). Subjects were prohibited from eating or drinking for 15 minutes before saliva collection and were asked to refrain from brushing their teeth or eating. Saliva was collected cryovially by drooling using Saliva Collection Aid (Salimetrics). The saliva samples were centrifuged (1500 x g) at 4°C for 10 min and stored in a -80°C freezer until analysis.

### Saliva collections and measurements of salivary cortisol concentrations

Cortisol, amylase, and chromogranin A concentrations were measured as stress markers in saliva samples. Salivary cortisol concentrations were measured using the YK241 Cortisol (Saliva) EIA kit (Yanaihara Institute, Shizuoka, Japan), salivary amylase concentrations using the SALIVARY alpha-AMYLASE KINETIC ENZYME ASSAY KIT (SALIMETRICS, PA, USA), and salivary chromogranin A concentration using the YK070 Human CgA EIA kit (Yanaihara Institute, Shizuoka, Japan) according to the manual. Salivary cortisol concentration was measured at 450/620 nm, salivary amylase concentration at 405 nm, and salivary chromogranin A concentration at 490 nm absorbance on a BioTek Synergy LX multimode plate reader (Agilent, CA, USA). A standard curve was prepared from the concentration and absorbance results of the standard solutions, and the concentration of each sample was calculated using a regression equation.

### Statistical analysis

The results of each measurement are expressed as mean or median values and the corresponding standard deviation (SD) or interquartile range. Multiple paired t-tests and the Wilcoxon matched-pairs signed rank test were used to identify statistically significant differences in the variables between the two videos. Statistical significance was set at p < 0.05. All statistical analyses were performed using Prism 9.5.1 (GraphPad Software, Boston, MA, USA).

### Ethics approval and consent to participate

Written informed consent was obtained from all participants. This study was approved by the Gunma University Medical Ethics Review Committee for Human Subjects (approval number: HS2020-147). All measurements on the participants were performed in accordance with the Declaration of Helsinki.

## Results

### The subjective score of the participants for each video

The subjective scores of the participants after viewing each video are shown in Fig 2 and Table 2. The scores after watching the high-interest videos are shown as median (25th –75th percentile): 6.50 [5.00–8.00] for concentration, 7.00 [5.25–7.00] for interest, and 8.00 [7.00–8.75] for comprehension. After watching the videos of low interest, the scores were 4.00 [3.00–5.00] for concentration, 2.50 [1.25–4.00] for interest, and 4.50 [3.00– 6.00] for comprehension. In both sets of scores, videos with high interest significantly exceeded those with low interest (p<0.0001). Conversely, the score for fatigue was 4.00 [2.25–5.75] after watching the videos of high interest, which was significantly lower than the score of 7.00 [5.00–8.00] after watching the videos of low interest (p<0.0001). Even when the results were examined separately for men and women, the scores for concentration, interest, and comprehension were significantly higher, and the scores for fatigue were substantially lower for videos of high interest than for videos of low interest. In particular, women showed a more pronounced difference in the subjective scores for high- and low-interest videos.

**Fig 2.**
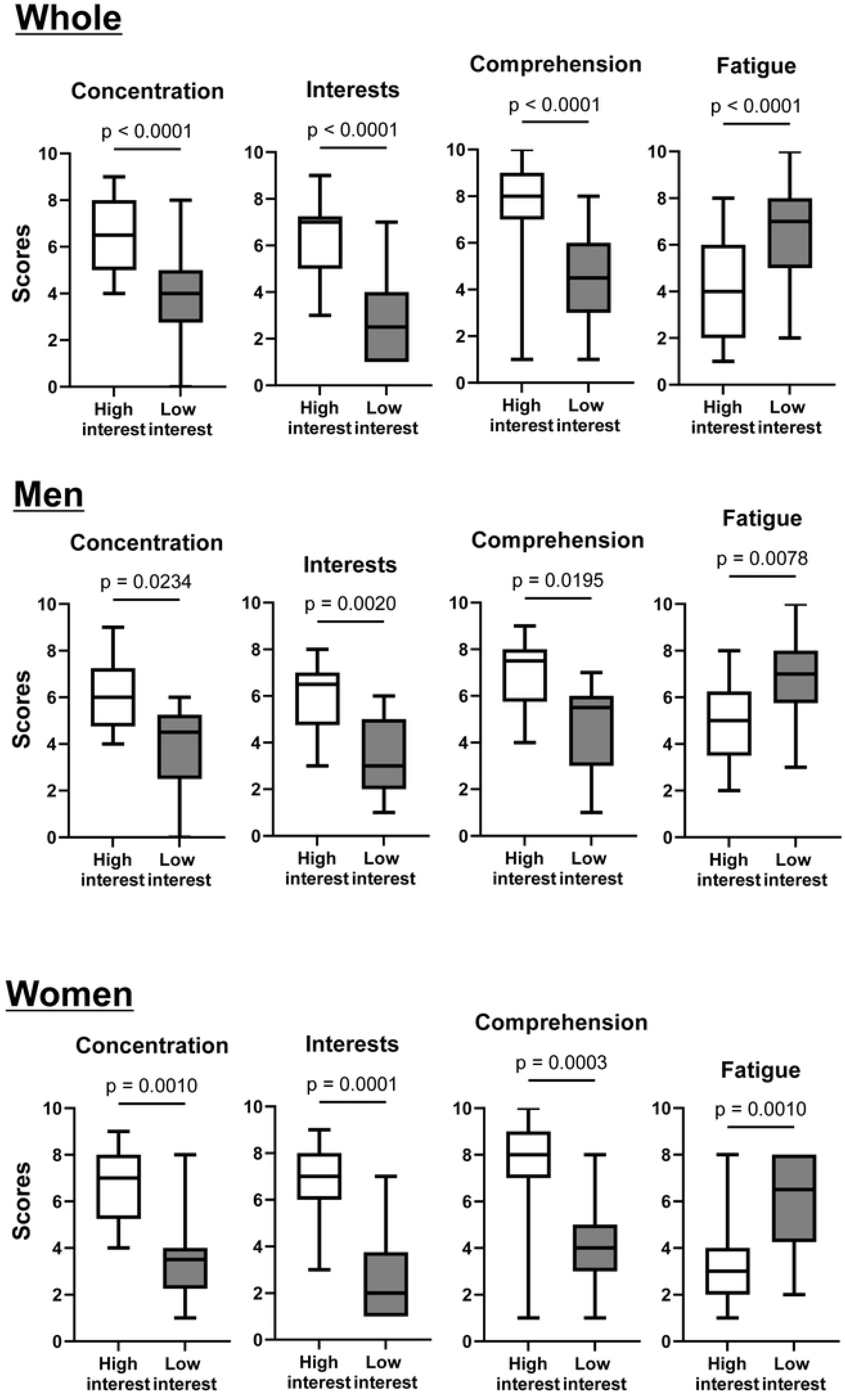
Comparison of the results of the VAS scale responses for concentration (A), comprehension (B), interest (C), and fatigue (D) after watching a video. White boxes indicate results after viewing videos of high interest, and gray boxes indicate results after viewing videos of low interest. Significant differences between the two videos were analyzed using the Wilcoxon signed-rank test.

**Table 1.** Characteristics of subjects.

| Characteristics | Values |
| --- | --- |
| Number | 26 |
| Age | 22.1 ± 0.7 |
| Male (%) | 10 (38.5) |

Data are expressed as mean ± SD or n (percentages).

**Table 2.** Comparison of subjective scores for watching high-interest and low-interest videos.

|  |  | High-interest | Low-interest | P value |
| --- | --- | --- | --- | --- |
| **Whole (n=26)** | Concentration | 6.50 (5.00-8.00) | 4.00 (3.00-5.00) | <0.0001 |
|  | Interests | 7.00 (5.25-7.00) | 2.50 (1.25-4.00) | <0.0001 |
|  | Comprehension | 8.00 (7.00-8.75) | 4.50 (3.00-6.00) | <0.0001 |
|  | Fatigue | 4.00 (2.25-5.75) | 7.00 (5.00-8.00) | <0.0001 |
| **Men (n=10)** | Concentration | 6.00 (5.00-7.00) | 4.50 (3.00-5.00) | 0.0234 |
|  | Interests | 6.50 (5.00-7.00) | 3.00 (2.00-4.75) | 0.0020 |
|  | Comprehension | 7.50 (6.25-8.00) | 5.50 (3.50-6.00) | 0.0195 |
|  | Fatigue | 5.00 (4.25-6.00) | 7.00 (6.00-8.00) | 0.0078 |
| **Women (n=16)** | Concentration | 7.00 (5.75-8.00) | 3.50 (2.75-4.00) | 0.0010 |
|  | Interests | 7.00 (6.00-8.00) | 2.00 (1.00-3.25) | 0.0001 |
|  | Comprehension | 8.00 (7.00-9.00) | 4.00 (3.00-5.00) | 0.0003 |
|  | Fatigue | 3.00 (2.00-4.00) | 6.50 (4.75-8.00) | 0.0010 |

Data are expressed as median (interquartile range). Significant differences between the high- and low-interest scores were analyzed using the Wilcoxon matched-pairs signed rank test.

### Heart Rate and Heart Rate Variability during Video Viewing

Fig 3 and Table 3 show the results of the time series of changes in heart rate, LF/HF, and HF during video viewing. Heart rate decreased rapidly during the 10 minutes after watching both types of videos compared to before viewing (p<0.0001). Compared with the heart rate when watching videos of low interest, the decrease in heart rate when watching videos of high interest was more pronounced and remained lower during all periods.

**Fig 3.**
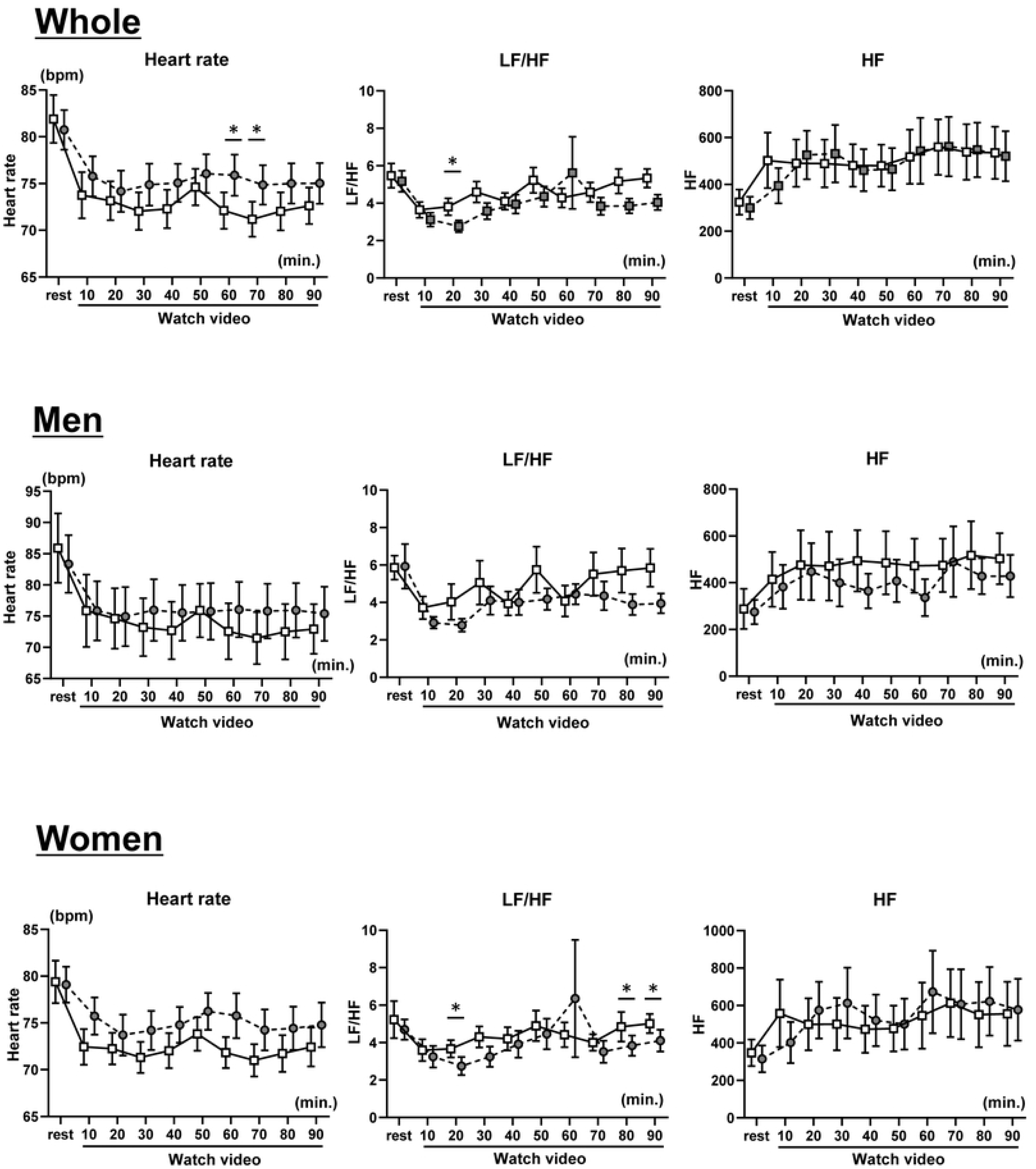
Comparison of heart rate (A), LF/HF (B), and HF (C) results during video viewing. White squares indicate results during video viewing of high interest, and gray circles indicate results during low interest. Data are shown as mean ± SEM. Significant differences between the two videos were analyzed using the multiple paired t-test.

**Table 3.** Comparison of heart rate variability for watching high-interest and low-interest videos.

|  |  | High-interest |  |  | Low-interest |  |  |
| --- | --- | --- | --- | --- | --- | --- | --- |
|  |  | pre | post | P value | pre | post | P value |
| **Whole (n=26)** | Heart rate (beat/min) | 81.9 ± 12.8 | 72.6 ± 9.8 | <0.0001 | 80.7 ± 10.6 | 75.0 ± 10.8 | <0.0001 |
|  | LF/HF | 5.47 ± 3.26 | 5.34 ± 2.48 | 0.8517 | 5.17 ± 2.82 | 4.05 ± 2.03 | 0.0807 |
|  | HF(msec<sup>2</sup>) | 324 ± 270 | 535 ± 557 | 0.0283 | 299 ± 237 | 520 ± 532 | 0.0098 |
| **Men (n=10)** | Heart rate (beat/min) | 85.9 ± 17.6 | 72.9 ± 12.6 | 0.0045 | 83.4 ± 14.6 | 75.4 ± 13.7 | 0.0281 |
|  | LF/HF | 5.86 ± 2.03 | 5.85 ± 3.20 | >0.9999 | 5.93 ± 3.77 | 3.95 ± 1.68 | 0.5176 |
|  | HF(msec<sup>2</sup>) | 288 ± 271 | 502 ± 343 | 0.0084 | 275 ± 163 | 429 ± 286 | 0.4606 |
| **Women (n=16)** | Heart rate (beat/min) | 79.4 ± 9.0 | 72.4 ± 8.3 | 0.0013 | 79.1 ± 7.7 | 74.8 ± 9.5 | 0.0025 |
|  | LF/HF | 5.23 ± 3.98 | 5.02 ± 2.05 | >0.9999 | 4.70 ± 2.16 | 4.11 ± 2.33 | 0.9398 |
|  | HF(msec<sup>2</sup>) | 348 ± 284 | 556 ± 683 | 0.6426 | 315 ± 285 | 577 ± 659 | 0.2059 |

Data are expressed as mean ± SD. Significant differences between the pre- and post-values were analyzed using the paired t-test.

As with heart rate, LF/HF values decreased more quickly at 10 minutes after watching the videos than at rest. For videos of high interest, the LF/HF values were significantly higher than those of low interest at 20 minutes but tended to approach the resting values after that. In the videos of low interest, the LF/HF values showed a decreasing trend up to 20 minutes after viewing but then showed an increasing trend. There were no significant differences in the LF/HF values before and after viewing the videos.

Contrary to the behavior of heart rate and LF/HF values, HF values increased quickly for up to 20 minutes after watching the videos. HF increased 10 minutes after viewing a video of high interest and was more pronounced than when viewing a video of low interest. Subsequently, the HF values remained at about the same level, and the HF values before and after viewing the videos increased significantly during both types of video viewing (p<0.05).

Results by sex showed that heart rate and LF/HF were similar to the overall results for men. HF was higher for most periods when viewing videos of high interest than when viewing videos of low interest. Although the heart rate for women showed similar trends to the overall results, the LF/HF showed significant increases at 20, 80, and 90 minutes post-viewing for videos of high interest compared to videos of low interest. HF increased after viewing all videos but showed higher values for videos of low interest, showing a different trend from that of men.

### Changes in Salivary Hormones during Video Viewing

Fig 4 and Table 4 show the changes in salivary cortisol, amylase, and CgA concentrations before, and at 45 and 90 minutes after viewing the videos. Salivary cortisol levels showed a significant decrease after viewing the videos compared to before viewing (p<0.0001). There was little difference in the extent of reduction between videos of high and low interest.

**Fig 4.**
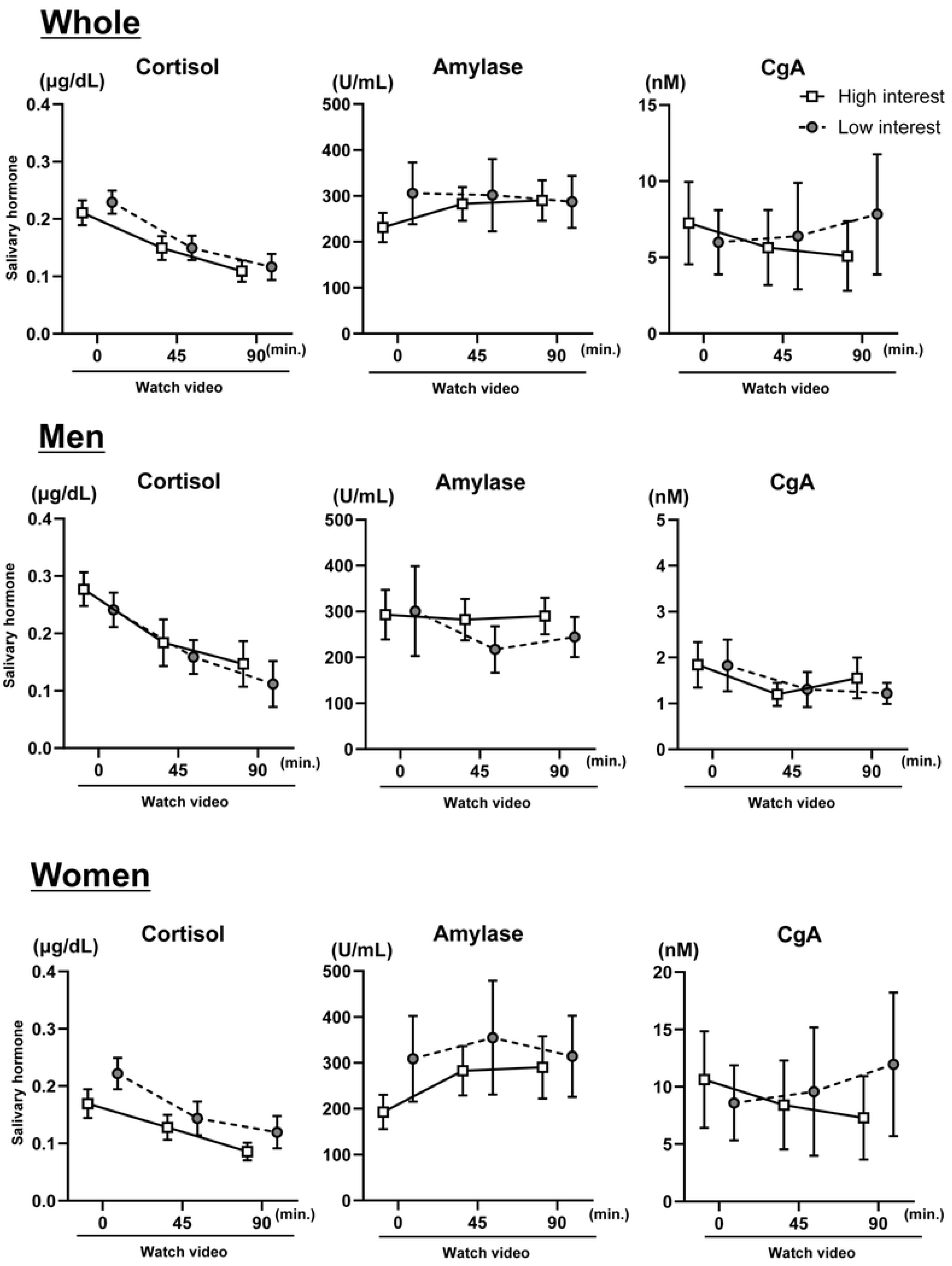
Comparison of salivary cortisol (A), salivary amylase (B), and salivary CgA (C) concentrations before, during, and after watching the videos. White squares indicate results for high interest during video viewing, and gray circles indicate results for low interest. Data are shown as mean ± SEM. Significant differences between the two videos were analyzed using the multiple paired t-test.

**Table 4.** Comparison of salivary hormones for watching high-interest and low-interest videos.

|  |  | High-interest |  |  | Low-interest |  |  |
| --- | --- | --- | --- | --- | --- | --- | --- |
|  |  | pre | post | P value | pre | post | P value |
| **Whole (n=26)** | Cortisol (mg/dL) | 0.211 ± 0.109 | 0.109 ± 0.094 | <0.0001 | 0.229 ± 0.102 | 0.117 ± 0.116 | <0.0001 |
|  | Amylase (U/mL) | 231 ± 162 | 290 ± 223 | 0.0706 | 305 ± 343 | 287 ± 288 | 0.5232 |
|  | CgA (pmol/mL) | 7.25 ± 13.80 | 5.08 ± 11.63 | 0.0112 | 5.99 ± 10.74 | 7.83 ± 20.10 | 0.4429 |
| **Men (n=10)** | Cortisol (mg/dL) | 0.277 ± 0.093 | 0.147 ± 0.126 | 0.0057 | 0.241 ± 0.095 | 0.112 ± 0.126 | 0.0069 |
|  | Amylase (U/mL) | 293 ± 171 | 290 ± 126 | 0.9942 | 301 ± 310 | 244 ± 138 | 0.6236 |
|  | CgA (pmol/mL) | 1.84 ± 1.56 | 1.55 ± 1.40 | 0.6091 | 1.83 ± 1.78 | 1.22 ± 0.72 | 0.3503 |
| **Women (n=16)** | Cortisol (mg/dL) | 0.170 ± 0.100 | 0.086 ± 0.061 | 0.0023 | 0.222 ± 0.109 | 0.120 ± 0.113 | 0.0029 |
|  | Amylase (U/mL) | 193 ± 150 | 290 ± 271 | 0.0764 | 310 ± 373 | 314 ± 354 | 0.9593 |
|  | CgA (pmol/mL) | 10.63 ± 16.85 | 7.29 ± 14.52 | 0.0243 | 8.60 ± 13.10 | 11.95 ± 25.02 | 0.5927 |

Data are expressed as mean ± SD. Significant differences between the pre- and post-values were analyzed using the paired t-test.

Salivary amylase levels increased slightly at 45 minutes after viewing a high-interest video compared to the resting state, but there was little difference until 90 minutes. There was almost no difference over time when viewing videos of low interest. Salivary amylase levels were not significantly different before and after watching the videos.

Salivary CgA concentration decreased at 45 and 90 minutes after viewing the high-interest video and was significantly reduced compared to that before viewing (p<0.0001). In contrast, salivary CgA concentrations tended to increase at 45 and 90 minutes after viewing videos of low interest. However, there was no significant difference in salivary CgA concentration after viewing compared to before viewing.

In the results examined for men and women, men showed trends similar to the overall results for salivary cortisol and amylase, with little difference between the two videos. Salivary CgA showed no change over time for either of the two videos, and the values were similar before and after video viewing. Women showed a decreasing trend in salivary cortisol before and after watching the videos, whereas salivary amylase showed an increasing trend. Both salivary cortisol and amylase levels were higher when viewing videos of low interest than when viewing videos of high interest. Salivary CgA showed a decreasing trend after viewing videos of high interest but increased after viewing videos of low interest.

## Discussion

In this study, we measured subjective scores, sympathetic and parasympathetic nervous system activity, and salivary stress hormone levels while participants watched two videos of differing levels of interest. In the subjective evaluation, concentration and comprehension were higher when watching the video of high interest, and fatigue was lower than when watching the video of low interest. In addition, heart rate decreased when watching each video, but this decrease was particularly pronounced when watching the high-interest video. Furthermore, LF/HF decreased, and HF tended to increase when watching each video; however, there was no difference between the videos. When comparing salivary hormones, cortisol decreased significantly after watching the video, whereas amylase showed almost no change. CgA decreased in videos that aroused a high level of interest, while it increased in videos that aroused a low level of interest. Analysis by sex showed that the difference in subjective scores between the high- and low-interest videos was more pronounced for women.

In addition, in women, the LF/HF of heart rate variability showed a significant increase in the high-interest video compared to the low-interest video, and salivary CgA decreased after watching the high-interest video and increased after watching the low-interest video. In contrast, there was almost no difference in heart rate variability or salivary hormone levels between the two videos in men.

### Watching videos affects heart rate, heart rate variability, and salivary hormones

In this study, heart rate decreased in both types of video viewing, with a decrease in LF/HF and an increase in HF of heart rate variability. As reported regarding video viewing and heart rate, in a comparison of 120 minutes of viewing a pleasant, unpleasant, and neutral movie, the heart rate decreased after watching all three types of videos, but the heart rate decreased more after watching the pleasant and unpleasant videos.[21] In addition, participants who viewed animal videos reported significantly lower heart rate and blood pressure levels and a reduced cardiovascular response to psychological stress compared to those who viewed human or blank-screen videos.[22] In the present study, neither video was mentally taxing enough to significantly affect the cardiovascular responses, which may have led to a decreased heart rate due to video viewing. It has also been reported that concentrating on mentally demanding tasks increases sympathetic nerve activity, increasing heart rate.[23] After 10 minutes of video viewing, LF/HF decreased, HF increased in both groups, and heart rate decreased during the same period, suggesting that heart rate decreased with the autonomic response.

In this study, salivary cortisol also decreased after viewing both videos. Salivary cortisol is associated with the HPA system and is known to increase in response to acute stress. Watching videos with aggressive content has increased testosterone and cortisol levels in saliva.[24] However, some reports suggest that physical stress, such as high-intensity exercise, also increases salivary testosterone and cortisol, and that cognitive stressors have no such effect,[25] suggesting that cortisol is more susceptible to physical and emotional stress. In this study, salivary cortisol also decreased with viewing time in both video groups, suggesting that salivary cortisol reflected a reduction in physical stress.

In contrast, salivary amylase remained unchanged, showing little change before and after viewing the two videos. It has been reported that salivary amylase levels decrease when viewing soothing videos, indicating its potential as a relaxation indicator.[26] In addition, salivary amylase activity increases upon exposure to emotional pictures, especially chopped pictures, suggesting a direct relationship between visual stimulus discomfort and amylase levels.[27] In this study, salivary amylase activity was altered by the emotional valence of the pictures but not by arousal ratings, an indicator related to discomfort induced by sustained exposure to emotional stimuli. In the current study, salivary amylase remained unchanged in both groups of videos, showing little change before and after viewing, suggesting that there was little emotional impact on the participants.

### The effect of differences in interest in watching videos on the physical and subjective scores

In this study, heart rate decreased in both groups during video viewing, but heart rate decreased more in the high-interest videos. HF increased quickly at 10 minutes after viewing the high-interest videos, while HF rose slowly in the low-interest videos, reaching the same level at 20 minutes after viewing. Sergii et al. reported that heart rate and parasympathetic nervous system activity decreased after viewing neutral and negative news, especially negative TV news.[28] However, the results from a random order of viewing stressful and neutral movies showed no difference in HRV between the two movies, although viewing the stressful movie resulted in an increase in heart rate.[29] In addition, it has been reported that watching 3D images can increase heart rate and the VLF/HF ratio, thus inducing sustained activation of the sympathetic nervous system, which can increase stress and emotional involvement.[30] The subjective scores for concentration, comprehension, and interest in the videos in this study suggest that the autonomic nervous system responds early when viewing videos of high interest, leading to an increase in parasympathetic nervous system activity and a decrease in heart rate. Furthermore, the subjective scores for fatigue suggest that psychological stress was reduced more in the high-interest videos and that there were differences in cardiovascular responses.

### Differences in physiological responses when watching videos between men and women

When the results were compared separately for men and women, the difference in the subjective scores for the two videos was particularly pronounced for women. In addition, heart rate decreased more when watching videos of high interest for women. HF increased more slowly when watching videos of low interest, reaching its peak 30 minutes after watching. Women generally show higher HF power than men and have more active parasympathetic nervous system activity.[31] However, under stress, it has been reported that women show a significant decrease in HF and that there are gender differences in stress-coping ability.[32, 33] The fact that the HF response to watching the two videos differed between women and men is thought to be one of the reasons for the difference in heart rate. In addition, research on sex differences in psychological stress and cardiovascular and sympathetic-adrenal responses has also reported that while men are characterized by increases in blood pressure and catecholamine responses, women are characterized by pronounced heart rate responses.[34] These data also suggest that a woman’s age and hormonal status can affect autonomic nervous system responses, and these factors were possibly involved in the decrease in heart rate observed in women in this study.

In addition, the concentration of salivary CgA showed different behaviors in men and women. Chromogranin A is associated with the secretion of catecholamines from the adrenal medulla and sympathetic nervous system. Salivary chromogranin A is known to correlate with increased sympathetic activity.[17] Salivary CgA has also been reported to decrease significantly after viewing a comedy video and increase immediately after completing a computational task.[35] In addition, one report shows that salivary CgA increases, but salivary cortisol does not change under the stress of a 60-minute written exam for nursing students, indicating its sensitivity as a short-term mental stress marker.[18] This study showed that while there was almost no change in the videos for both groups of men, there was a decrease in the videos of high interest for women and an increase in the videos of low interest, suggesting that this is a reaction unique to women. Previous reports have shown no baseline differences between men and women or differences in stress responses for salivary CgA.[36] This difference in the stress response of salivary CgA may be related to the menstrual cycle, hormonal influences, and the degree of stress experienced by individuals. It has also been reported that psychological stress factors can cause gender differences, with men showing a higher salivary hormone response to achievement-oriented tasks and women showing a stronger response to social rejection.[37] The subjective scores for the current video also showed more significant differences in women; therefore, the type of video possibly affected the differences in salivary CgA.

This study showed a difference in reactivity between the two videos for female HF, with a gradual increase and decrease in reactivity in the low-interest video. Salivary CgA is a substance related to the SAM system and is closely related to autonomic nervous system activity. However, many aspects, such as the relationship between heart rate variability and differences in reactivity depending on the level of psychological stress, have not yet been clarified. Further investigation of the relationship between stress by combining heart rate variability and salivary stress markers is necessary.

### Limitations of the Study

Our study has several limitations. First, although we used two types of videos—one with high interest and one with low interest—the same video was used for all subjects; therefore, interest was not necessarily matched. Some participants who were prepared for videos of low interest responded as highly interested on the VAS scale. Further research is needed to determine whether our findings are specific to the videos used in this study or whether they can be generalized to other videos with similar content.

Second, because saliva samples were collected at three separate time points, we were unable to analyze the interrelationship between salivary stress hormones and variations in the autonomic nervous system. Salivary cortisol and amylase are considered to have a time lag after stress,[20] making it difficult to analyze their interrelationships with autonomic nervous system variations in real-time. Future research should include methods such as simultaneous sampling and analysis.

## Conclusions

This study demonstrated that viewing videos of high and low interest for extended periods affected autonomic nervous system activity, heart rate, and salivary stress hormones. The difference in subjective scores was particularly pronounced in women, and differences were also seen in the parasympathetic nervous system and salivary stress hormones.

Since the effects on the autonomic nervous system and stress hormones are more significant when people repeatedly watch videos for long periods, such as during remote lectures and meetings, the combination of autonomic analysis and salivary stress hormone analyses may help assess adjustment disorders and excessive stress.

## Data Availability

All relevant data are within the manuscript and its Supporting Information files.

## Acknowledgments

We would like to thank all the participants who contributed to this study.

## Notes

### Competing Interest Statement

The authors have declared no competing interest.

### Funding Statement

Yes

### Author Declarations

This study was approved by the Gunma University Medical Ethics Review Committee for Human Subjects (approval number: HS2020-147).

